# Routine asymptomatic testing strategies for airline travel during the COVID-19 pandemic: a simulation analysis

**DOI:** 10.1101/2020.12.08.20246132

**Authors:** Mathew V Kiang, Elizabeth T Chin, Benjamin Q Huynh, Lloyd A C Chapman, Isabel Rodríguez-Barraquer, Bryan Greenhouse, George W Rutherford, Kirsten Bibbins-Domingo, Diane Havlir, Sanjay Basu, Nathan C Lo

## Abstract

**Background:** Airline travel has been significantly reduced during the COVID-19 pandemic due to concern for individual risk of SARS-CoV-2 infection and population-level transmission risk from importation. Routine viral testing strategies for COVID-19 may facilitate safe airline travel through reduction of individual and/or population-level risk, although the effectiveness and optimal design of these “test-and-travel” strategies remain unclear.

**Methods:** We developed a microsimulation of SARS-CoV-2 transmission in a cohort of airline travelers to evaluate the effectiveness of various testing strategies to reduce individual risk of infection and population-level risk of transmission. We evaluated five testing strategies in asymptomatic passengers: i) anterior nasal polymerase chain reaction (PCR) within 3 days of departure; ii) PCR within 3 days of departure and PCR 5 days after arrival; iii) rapid antigen test on the day of travel (assuming 90% of the sensitivity of PCR during active infection); iv) rapid antigen test on the day of travel and PCR 5 days after arrival; and v) PCR within 3 days of arrival alone. The travel period was defined as three days prior to the day of travel and two weeks following the day of travel, and we assumed passengers followed guidance on mask wearing during this period. The primary study outcome was cumulative number of infectious days in the cohort over the travel period (population-level transmission risk); the secondary outcome was the proportion of infectious persons detected on the day of travel (individual-level risk of infection). Sensitivity analyses were conducted.

**Findings:** Assuming a community SARS-CoV-2 incidence of 50 daily infections, we estimated that in a cohort of 100,000 airline travelers followed over the travel period, there would be a total of 2,796 (95% UI: 2,031, 4,336) infectious days with 229 (95% UI: 170, 336) actively infectious passengers on the day of travel. The pre-travel PCR test (within 3 days prior to departure) reduced the number of infectious days by 35% (95% UI: 27, 42) and identified 88% (95% UI: 76, 94) of the actively infectious travelers on the day of flight; the addition of PCR 5 days after arrival reduced the number of infectious days by 79% (95% UI: 71, 84). The rapid antigen test on the day of travel reduced the number of infectious days by 32% (95% UI: 25, 39) and identified 87% (95% UI: 81, 92) of the actively infectious travelers; the addition of PCR 5 days after arrival reduced the number of infectious days by 70% (95% UI: 65, 75). The post-travel PCR test alone (within 3 days of landing) reduced the number of infectious days by 42% (95% UI: 31, 51). The ratio of true positives to false positives varied with the incidence of infection. The overall study conclusions were robust in sensitivity analysis.

**Interpretation:** Routine asymptomatic testing for COVID-19 prior to travel can be an effective strategy to reduce individual risk of COVID-19 infection during travel, although post-travel testing with abbreviated quarantine is likely needed to reduce population-level transmission due to importation of infection when traveling from a high to low incidence setting.

## Introduction

The COVID-19 pandemic has dramatically changed daily life for people and reduced global travel.^1^ Since the first report of a novel coronavirus in Wuhan, China on December 31, 2019, the causative virus SARS-CoV-2 has spread globally with an unprecedented number of cases and deaths.^2^ The principal public health strategy has been the implementation of universal masking and social distancing interventions aimed at reducing the number of social interactions to slow the spread of the virus.^3-5^ The approach of social distancing has effectively resulted in the closure of businesses, workplaces, schools, and other public venues. Due to these recommendations and closure of national borders in an attempt to curb the spread, domestic and international airline travel has been dramatically reduced by over 80% in the year following the report of the original outbreak.^6-8^ The decrease in airflight travel is likely due to multiple etiologies including personal choice to minimize the risk of infection, governmental and employer policy enforcing travel restrictions or quarantine requirements, cancellation of professional and social events requiring travel, and other motivations for reducing travel.^6,7^

Asymptomatic viral testing strategies for SARS-CoV-2 may facilitate safe airline travel through reduction of individual risk of infection and/or population-level risk from importation of infection related to travel. Up to 50% of persons with SARS-CoV-2 infection are asymptomatic and do not know about their infection, and this population contributes a large proportion of new cases and transmission.^9^ The strategy of routine viral testing during travel has two possible applications: i) reduction in individual risk of infection in the airport/airplanes by detecting infected passengers and preventing their travel; and ii) reduction in the number of importations of infections to a new city, hence reducing the impact of travel on population-level transmission risk. Currently, the mainstay strategy has been to avoid traveling altogether, although this is likely to change over time. The majority of persons in the United States who travel do so without testing or self-isolation requirements. Some travelers may elect for testing prior to travel or upon arrival, and in some cases airlines have begun to offer testing for SARS-CoV-2 prior to departure.^10^ In other cases, travelers may elect or be required to have a self-isolation period upon arrival in the absence of testing.^11^ Recently, the United States Centers for Disease Control and Prevention (CDC) released recommendations for asymptomatic testing for COVID-19 including getting tested 1-3 days before the flight, and tested 3-5 days after travel with a post-travel quarantine period.^12^

There are many considerations for the design of routine testing strategies to minimize individual and/or population risk of COVID-19 associated with travel. Testing strategies designed to minimize individual-level risk will focus on pre-travel testing, while strategies designed to reduce population-level transmission will include both pre- and post-travel testing. The choices of test can include: i) polymerase chain reaction (PCR), the current diagnostic gold-standard, with very high sensitivity but slow turnaround time; or ii) rapid tests (either antigen or nucleic acid based) that are becoming increasingly available, have fast turnaround time (< 30 mins) and have been shown to have good (although variable between manufacturers) sensitivity to detect infection during their most infectious period.

Domestic and international airline travel is likely to rise over time, although the optimal approach for a “test-and-travel” strategy and its effectiveness to minimize the risk of COVID-19 during airline travel remains unclear. The goal of this study was to identify the most effective “test-and-travel” protocol alongside universal masking to reduce the spread of COVID-19.

## Methods

### Model structure

We extended a previously published microsimulation model of SARS-CoV-2 transmission to identify the optimal testing strategy to detect infectious airline travelers before travel or soon after arrival to their destination.^13^ We simulated a population of 100,000 individual United States domestic airline travelers over their travel period, including three days prior to travel (when the earliest testing would take place), the day of travel, and two weeks after arrival at their final destination. This time horizon was chosen to fully capture the impact of all testing strategies on both individual and population risk upon arrival. Each traveler was assigned a single health state at a given point in time from one of five states that included: being susceptible to infection (non-immune), incubation period, early infectious (the pre-symptomatic infectious period), late infectious (symptomatic infectious period, if symptomatic), or recovered (with immunity). We assumed a mean infectious duration of five days.^9,14^ We used a static infection model, meaning each individual had a fixed probability of being infected each day, with a modestly increased risk during the travel period.^7,15,16^ We estimated a per-day risk of infection with SARS-CoV-2 corresponding to a daily incidence of 50 infections per 100,000 persons, which is representative of the current incidence observed in many US states as of December 1, 2020 (Supplemental Materials).^17^ The daily risk of infection was varied in sensitivity analysis from 1 to 250 daily infections per 100,000 persons. We simulated new infections starting two months prior to travel and three days days after travel to simulate transmission around the travel period; we counted the number of infectious days over the travel period, defined as three days before travel to two weeks after travel to include any infections related to travel. We assumed a 2-fold increased risk of infection on the day of travel, which is an assumption based on the number of social interactions in the airport/airplane and published literature, including with airport employees without daily testing.^7,15,16^ This assumption was varied in sensitivity analysis, including a scenario with no increased risk in the airport due to screening, ventilation, social distancing, and masking.^7,15,16^ We did not explicitly account for differences in duration of air-flight travel given paucity of literature to inform differential risk, and modeled only one-way travel. We accounted for a number of unique features of the natural history of COVID-19, including the incubation and infectious periods, proportion of sub-clinical (often asymptomatic) infection, pre-symptomatic transmission, and severity of illness (see Table 1).^14,18,19^ We did not include a baseline recovered fraction in the population in the model given low seroprevalence estimates in the general population.^20^ More details on the model structure and parameters are available in the Supplemenetal Materials, published code, and prior publication.^13,21^

**Table 1:**
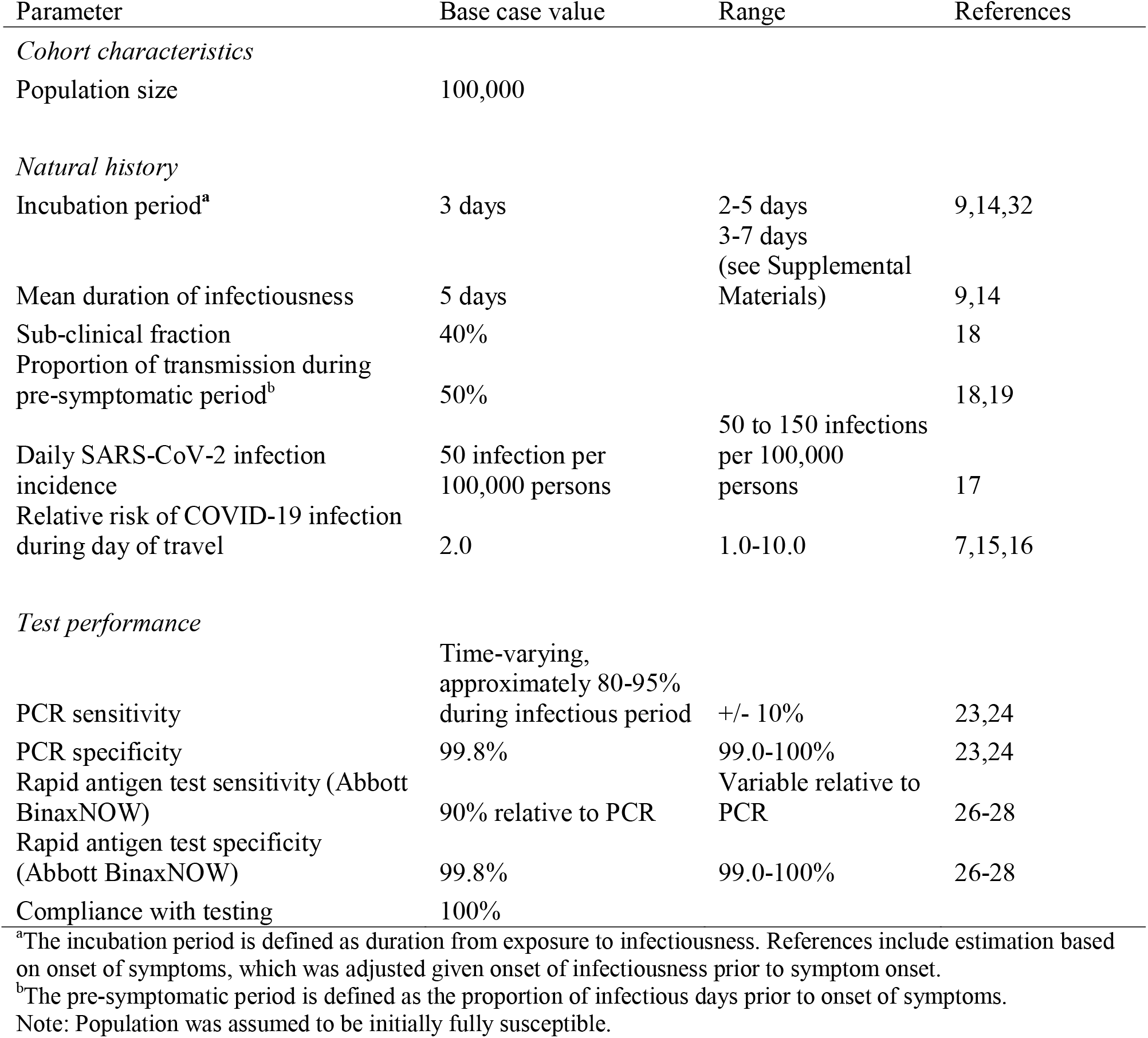
Cohort characteristics and COVID-19 natural history and transmission parameters.

### Simulation of testing strategies

We evaluated five viral testing strategies of airline travelers around the time of travel, including: i) anterior nasal PCR within 3 days of departure, meaning passengers were tested 2-3 days prior to the day of travel; ii) PCR within 3 days of departure and PCR on day 5 after arrival with 5 days of quarantine upon arrival; iii) rapid antigen test on the day of travel; iv) rapid antigen test on the day of travel and PCR on day 5 after arrival with 5 days of quarantine upon arrival; and v) PCR within 3 days of arrival, meaning passengers were tested 1-2 days after arrival. These strategies were chosen based on informal consultation with experts, recent guidance from the United States Centers for Disease Control and Prevention,^22^ and state policies. The focus of post-travel testing and quarantine was to reduce importation of infections acquired immediately prior to departure. Notably, the testing strategies differentially affect the individual risk of infection while traveling (strategies i-iv) versus decreasing population-level transmission risk from importation of new infections to a destination city (strategies ii, iv, and v). We assumed that symptomatic persons avoided travel with perfect compliance. We assumed that travelers who tested positive did not travel, and self-isolated with perfect compliance. We did not include any self-quarantine requirements upon arrival in the destination for pre-travel-only testing strategies (i and iii). We used published literature on the sensitivity and specificity of rapid antigen tests and PCR assays for SARS-CoV-2, incorporating time-varying estimates of sensitivity based on time since exposure.^23^ PCR tests were assumed to have sensitivity ranging from 80-95% during the first two weeks following exposure with a peak in sensitivity by day five based on the upper limit estimate on sensitivity from a published study (following the viral kinetics over time within individuals; see Supplemental Materials for curve of test sensitivity over time)^23^, and specificity of 99.5-100% (mean 99.8%).^23,24^ We assumed a one-day turnaround time for PCR tests. Persons were assumed to be detectable by PCR for up to two weeks after no longer being infectious. ^23^ For rapid antigen tests, there is wide variation in the sensitivity and specificity of these assays. We assumed the rapid antigen tests had the sensitivity and specificity of the Abbot BinaxNow based on availability of literature to support a sensitivity of 90% relative to PCR (during periods of high viral loads compatible with infectiousness) and specificity of 99.5-100% (mean 99.8%). Other rapid testing platforms have variable sensitivity and specificity (including some <50% sensitivity compared to PCR), although some may have comparable specification.^25-28^ The 95% uncertainty intervals (UI) represent the 2.5^th^ to 97.5^th^ percentile values across all simulations. The data and code are available online.^21^

### Study outcomes

The primary study outcome for each testing strategy was the cumulative number of infectious days in the cohort of airline travelers prior to detection of infection and self-isolation over the travel period.^14^ This outcome is most relevant for population-level risk of transmission, and was selected based on the public health goal of reducing overall transmission. The secondary outcomes included: i) proportion of infectious travelers with SARS-CoV-2 detected on the day of travel, which is most relevant for individual risk of infection during travel and from the airlines whose goal is to minimize transmission at the airport and during air flight travel; and ii) the ratio of false positives to true positives for each testing strategy and infection incidence.

### Sensitivity analysis

We performed sensitivity analyses to measure the effect of varying individual and multiple model parameters on the overall study conclusion. We tested varying individual model inputs including natural history parameters for SARS-CoV-2-such as duration of infectiousness, test sensitivity and specificity, day-of-travel increase in risk, and daily infection incidence.

## Results

### Descriptive results

In a cohort of 100,000 airline travelers, the model predicted 229 (95% UI: 170, 336) actively infectious persons on the day of travel in the absence of testing or any symptom screening (prevalence of 0.2% of actively infectious persons). Of these, 91 (95% UI: 63, 136) had subclinical infections. Over the travel period (three days prior to travel to two weeks following travel), we estimated a total of 2,796 (95% UI: 2,031, 4,336) infectious days.

### Effectiveness of test-and-travel strategies

First, we estimated that anterior nasal PCR testing within 3 days of departure would be positive for 1,001 travelers (95% UI: 817, 1,205), reducing the number of actively infectious travelers on the day of travel by 88% (95% UI: 76, 94) to 28 (95% UI: 13, 73) (see Table 2). This testing strategy reduced the total number of infectious days in the cohort by 35% (95% UI: 27, 42) over the simulation period (see Figure 1). At the assumed incidence of 50 daily infection per 100,000 and test specificity of 99.8%, there were 205 (95% UI: 31, 397) false positives. The ratio of false positives to true positives for this strategy ranged from 2.6 to 0.13 when varying incidence from 5 to 100 daily infections per 100,000 persons, respectively (see Figure 2).

**Table 2:** Effectiveness of test-and-travel strategies on study outcomes.

| Strategy | Number of infectious days | Study outcomes |  |
| --- | --- | --- | --- |
|  |  | Relative reduction in active infections on day of flight | Ratio of false positives to true positives |
| No testing, no screening | 2,796 (2,031, 4,336) | — | — |
| PCR test within 3 days of departure | 1,810 (1,274, 2,899) | 0.88 (0.76, 0.94) | 0.26 (0.04, 0.50) |
| PCR test within 3 days of departure;<br>PCR test within 5 days after arrival<sup>a</sup> | 599 (416, 1,072) | 0.88 (0.76, 0.94) | 0.38 (0.06, 0.74) |
| Rapid antigen testing on day of travel | 1,896 (1,352, 2,992) | 0.87 (0.81, 0.92) | 0.50 (0.08, 0.98) |
| Rapid antigen testing on day of travel;<br>PCR test within 5 days after arrival<sup>a</sup> | 824 (603, 1,300) | 0.87 (0.81, 0.92) | 0.65 (0.10, 1.28) |
| PCR test within 3 days after arrival | 1,630 (1,103, 2,766) | — | 0.38 (0.06, 0.74) |

Note: All testing strategies assume no symptomatic passengers will travel.
<sup>a</sup>These strategies include a 5-day quarantine period upon arrival.

**Figure 1:**
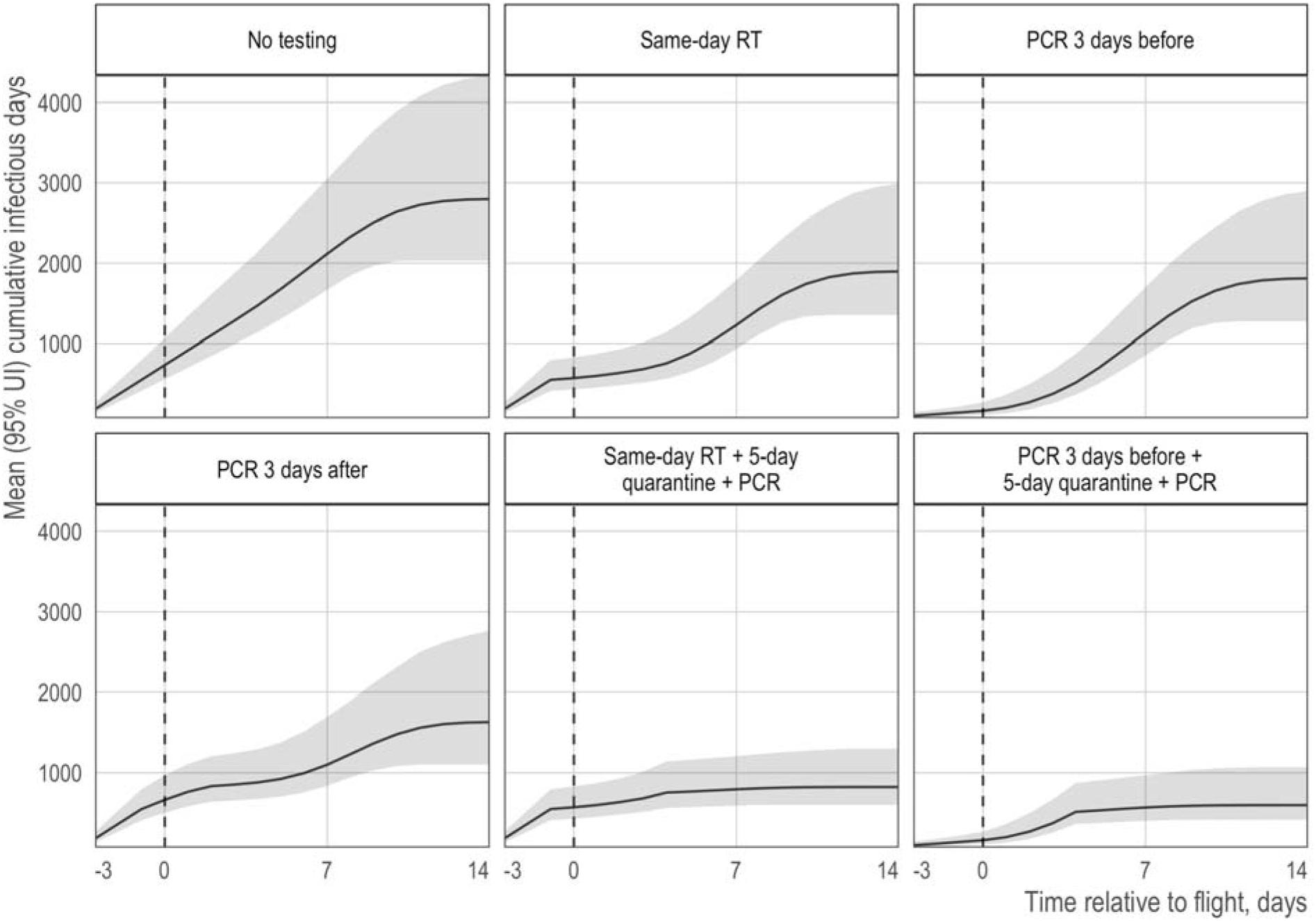
Predicted number of cumulative SARS-CoV-2 infectious days over a simulation period under different test-and-travel strategies. We estimated the number of infectious days (vertical axis) over time with simulation of each test-and-travel strategy (panels). The horizontal axis represents the time over the simulation (in days) relative to the day of travel (vertical dashed line). Solid lines represent the mean and shaded areas represent the 95% uncertainty interval (95% UI) across 3,000 simulations. RT, rapid antigen test; PCR, polymerase chain reaction.

**Figure 2:**
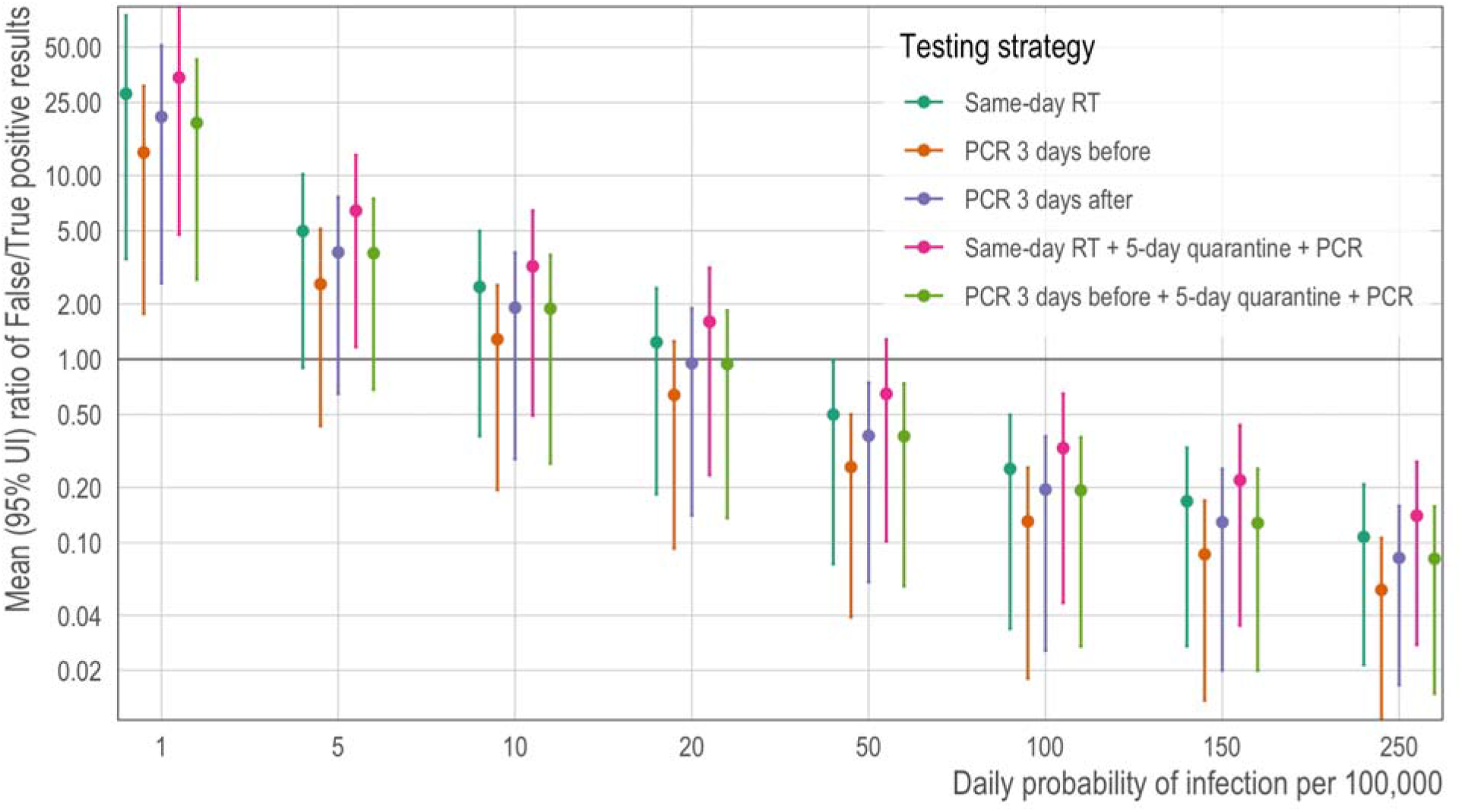
Ratio of false positive to true positive test results for test-and-travel strategies under different baseline SARS-CoV-2 infection incidence. We ran the microsimulation under different baseline SARS-CoV-2 infection incidence to understand the effect on ratio of false positives to true positives for each test-and-travel strategy. The horizontal axis represents SARS-CoV-2 infection incidence, including asymptomatic cases (daily cases per 100,000 persons). The vertical axis represents the ratio of false positives to true positives, where a higher ratio corresponds to a higher number of false positives. The colors of the lines correspond to the different test-and-travel strategies.

Second, we evaluated the strategy of anterior nasal PCR testing within 3 days of departure and PCR testing 5 days after arrival with quarantine for 5 days. This strategy yielded 1,489 positive travelers ((95% UI: 1,130, 1,886); defined as having any positive test), and reduced the number of actively infectious travelers on the day of travel by 88% (95% UI: 76, 94) to 28 travelers (95% UI: 13, 73). This testing strategy reduced the total number of infectious days in the cohort by 79% (95% UI: 71, 84) over the simulation period, with approximately 74% attributable to the pre-travel PCR and 26% attributable to the post-travel PCR test. The ratio of false positives to true positives for this strategy ranged from 3.8 to 0.19 when varying incidence from 5 to 100 daily infections per 100,000 persons, respectively.

Third, we estimated that the rapid antigen test on the day of travel would be positive for 618 passengers (95% UI: 445, 815), reducing the number of actively infectious travelers by 87% (95% UI: 81, 92) to 30 travelers (95% UI: 17, 51) on the day of travel. This testing strategy reduced the total number of infectious days in the cohort by 32% (95% UI: 25, 39) over the simulation period. There were 205 (95% UI: 31, 401) false positives, corresponding to a ratio of false to true positives that ranged from 5.0 to 0.25 when varying incidence from 5 to 100 daily infections per 100,000 persons, respectively.

Fourth, we evaluated a rapid antigen test on the day of travel and PCR testing 5 days after arrival with quarantine for 5 days. This strategy yielded 1,047 positive travelers ((95% UI: 686, 1438); defined as having any positive test). On the day of travel, the rapid antigen test would reduce the number of actively infectious travelers by 87% (95% UI: 81, 92) to 30 passengers (95% UI: 17, 51). This testing strategy reduced the total number of infectious days in the cohort by 70% (95% UI: 65, 75) over the simulation period, with approximately 65% attributable to the rapid antigen test and 35% attributable to the post-travel PCR test. The ratio of false positives to true positives for this strategy ranged from 6.4 to 0.32 when varying incidence from 5 to 100 daily infections per 100,000 persons, respectively.

Fifth, we estimated that anterior nasal PCR testing 3 days after arrival would be positive for 740 travelers (95% UI: 562, 941). This testing strategy reduced the total number of infectious days in the cohort by 30% (95% UI: 19, 42) over the simulation period. There were 205 (95% UI: 33, 398) false positives. The ratio of false positives to true positives for this strategy ranged from 3.8 to 0.19 when varying incidence from 5 to 100 daily infections per 100,000 persons, respectively.

### Sensitivity analysis

In sensitivity analysis, we varied single model inputs to determine their impact on study results. We found that lowering the sensitivity of tests reduced the proportion of cases detected and the number of infectious days averted, while lower specificity of tests significantly increased the number of positive tests and proportion of false positives. As long as the rapid antigen test sensitivity was greater than 90% compared to PCR, we found that the same-day rapid test performed similar to pre-travel PCR on detection of infectious passengers (see Supplemental Materials). A higher incidence of COVID-19 increased the absolute effectiveness of all testing strategies with a higher number of averted infectious days (see Figure 3). In a destination city with a higher incidence (250 daily infections per 100,000 persons), the post-travel testing provided modest relative reduction of total COVID-19 cases; in contrast, a destination city with a low incidence (10 daily infections per 100,000 persons), the post-travel testing provided a large relative reduction of cases (see Supplemental Materials).

**Figure 3:**
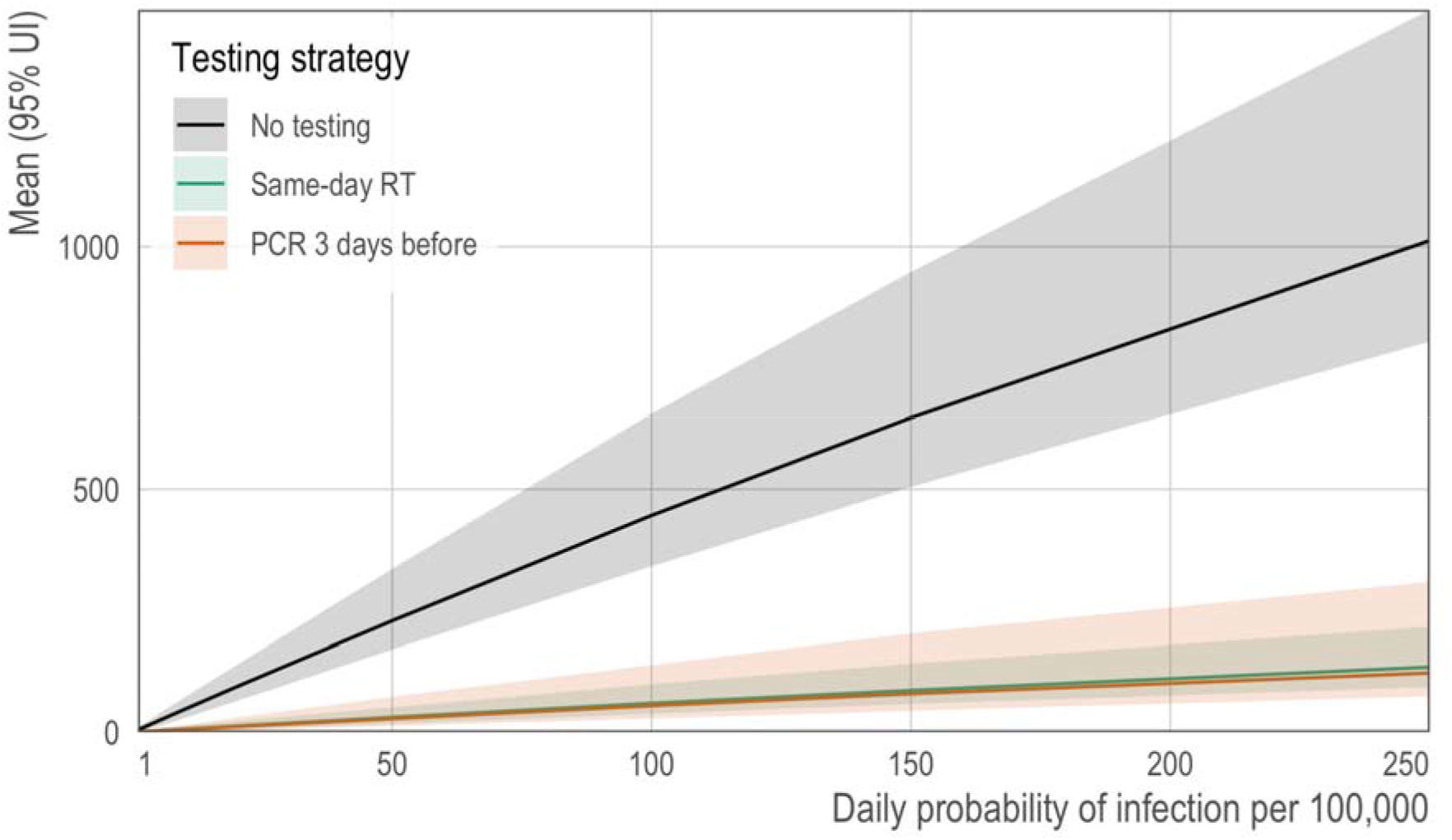
Effect of pre-travel testing strategies on the absolute number of travelers with SARS-CoV-2 infection. We estimated the mean number of total actively infectious persons on the day of travel in the cohort of 100,000 (vertical axis) under each pre-travel testing strategy: the base case (no testing), PCR within 3 days of departure, and rapid antigen test on the day of travel. We varied the baseline SARS-CoV-2 infection incidence (horizontal axis) from 1 to 250 daily infections per 100,000 persons. The vertical axis represents the mean and 95% uncertainty interval (95% UI) across 3,000 simulations. RT, rapid antigen test; PCR, polymerase chain reaction.

## Discussion

This simulation study finds that “test-and-travel” strategies for COVID-19 that apply routine viral testing during airline travel can reduce both the individual and population-level transmission risk of COVID-19 during travel. We find that pre-travel testing with a rapid antigen test on the day of travel or PCR testing within 3 days prior to departure can both reduce the risk of COVID-19 transmission during travel, with the majority of benefit for other travelers that may otherwise have become infected. We find that the addition of post-travel testing and abbreviated quarantine further provides further benefit at the level of the public health by reducing importation and ongoing transmission in the destination city, especially if travling from high to low incidence setting. Given that the prevalence of infection was relatively low, asymptomatic testing strategies resulted in a relatively high relative number of false positives due to imperfect test specificity (99.8%); this strategy also detected individuals that were test positive but no longer infectious (most relevant for PCR). Overall, this study supports that a test-and-travel strategy for COVID-19 will likely improve the safety of airline travel and can incorporated into national policy as a public health tool during the COVID-19 pandemic, alongside social distancing, universal masking, and other infection control measures during travel.

We found that all test-and-travel strategies had some benefit and each had strengths and drawbacks. The rapid antigen test had the advantage of timing since it can be administered on the day of travel with immediate turnaround, meaning the test is optimally timed to detect an infectious individual prior to flying. Notably, currently available rapid tests (antigen and nucleic acid-based) have highly variable test sensitivity and specificity. In this study, we parameterized the model focusing on the Abbott BinoxNOW or comparable tests that demonstrate reasonably high sensitivity (90% realative to PCR during the infectious period), with peak sensitivity based on the viral dynamics during the actively infectious period.^25-27^ Alternative rapid testing platforms such as loop-mediated isothermal amplification (LAMP) assays with similar test characteristics to the rapid antigen tests are also viable options if available as point-of-care tests.^25^ The PCR test within 3 days of travel had the benefit of higher analytic sensitivity as PCR remains the gold standard diagnostic, but has the drawback of slower turnaround time, can be less convenient, and has ongoing detection of prior SARS-CoV-2 infections that are no longer infectious. Given the delay in turnaround, this means the PCR tests are likely to be performed in the days prior to travel, and may miss an individual who is not yet exposed to COVID-19 or is exposed but has not yet become positive by viral testing, which usually occurs around 3-5 days after exposure. Both strategies appeared to have similiar benefit for reducing the number of infectious travelers although same-day rapid antigen tests are not currently recommended by the US CDC guidelines.^12^ We also examined the addition of a post-travel testing strategy with an abbreviated 5-day quarantine period, which notably performed much better on the study outcome of reducing overall infectious days associated with travel.

There are two key perspectives to interpret the study results that affect the conclusions drawn. The public health perspective (e.g. destination city of travel) would naturally focus on reducing population-level transmission with a goal of reducing importation of cases and ongoing transmission, and the primary outcome of reduction in infectious days would be most relevant. In this case, the testing strategies that include post-travel quarantine and PCR testing appear more favorable, especially when traveling from high to low incidence setting where importation of infection poses significant public health risk, and to a lesser extent, those infected during travel itself. Strategies with only pre-travel testing performed worse on the outcome of reducing total infectious days. In contrast, the focus of airlines is to reduce individual-level risk during travel as measured by the secondary outcome of the proportion of infectious travelers detected. In this case, the pre-travel testing strategies, especially rapid antigen test on day of travel, appear most favorable and efficient. However, some actively infected travelers were missed, this was mostly related to imperfect test sensitivity and persons who were exposed during travel but not yet detectable during the time of testing (in the case of PCR testing 2-3 days prior to travel). Further refinements could be made with consideration of COVID-19 incidence in the origin and destination cities to influence testing requirements, such as traveling from a high to low incidence city. In the case of pre-travel PCR testing, recommendations for strict social distancing in the days prior to travel would enhance the effectiveness of this strategy. Notably all strategies missed a sizable proportion of COVID-19 infections, and test-and-travel strategies overall should be viewed as a risk mitigation control strategy, which should be adopted in combination with social distancing, universal masking, and other infection control measures during airline travel.^3,15^

While the test-and-travel strategies were able to identify a large number of the actively infectious travelers, the testing strategies also had a large number of positive tests in travelers who were not infectious. This included: i) false positives due both to imperfect specificity of the test; and ii) persons who were previously infected and will test positive for up to 2-4 weeks after exposure but are no longer infectious (most relevant for PCR).^14^ The implication of this result means that some passengers will be unable to travel despite not being infectious or a risk to other passengers, but this is consistent with some international travel requirements. Airlines could consider additional testing when a false positive is suspected, such as when individuals are asymptomatic and have no known exposures; this confirmation testing could include a second rapid test (preferably on a different platform without similar false positive characteristics) or more extensive testing including serial PCR tests and later serologic test that may take multiple days and require self-isolation during this period (likely occurring outside of the airline testing system).

The study conclusions are supported by studies that have examined the effectiveness of asymptomatic testing strategies for COVID-19 to detect cases and reduce overall transmission.^13,29-31^ This literature has found that frequent testing, rapid turnaround of test results (within 1-2 days), and test sensitivity are key characteristics for an effective testing platform.^13,29,30^ In our sensitivity analysis in this study, we similarly found that delayed turnaround time or lower sensitivity significantly reduced the effectiveness of test-and-travel strategies. The asymptomatic testing strategies outside of the travel context are often designed as control strategies, meaning the goal is to reduce the risk of transmission but not detect all cases. The key evidence gap that the present study aimed to address was the application of asymptomatic testing to airline travel, as domestic and international air travel begins to increase over time.

The study findings should be interpreted within the context of the limitations of the data and model assumptions. There remain key uncertainties in the natural history of COVID-19, heterogeneity in transmission, and variation in diagnostic test accuracy and quality that are simplified in this modeling analysis. We assume a high degree of participation in testing programs and perfect self-isolation of those testing positive. We further assume that persons that are symptomatic do not travel based on current guidance, and the testing strategies would perform better if this was not true. The model assumes a fixed incidence and homogenous risk of infection for all persons, although we explore how different infection risk would affect our results. We include a modestly higher risk of infection during the day of airflight travel, although this risk is uncertain and assumed to be relatively low.^7,15,16^ We assumed all flights had comparable risk, despite variation in number of passengers and duration of flight. Routine asymptomatic test-and-travel strategies would require considerable resources, coordination, guidelines, and ongoing participation for airlines and travelers.

In conclusion, this study supports adopting asymptomatic testing strategies for COVID-19 in airline passengers to reduce the risk of travel and transmission during the pandemic. Multiple approaches to asymptomatic test-and-travel are likely to have benefit, although each have drawbacks that should be considered, and they will not reliably prevent transmission or outbreaks. This study provides evidence to guide national policy on testing guidelines to reduce COVID-19 spread from air travel. Continued efforts to promote social distancing, universal masking, and reducing travel will be necessary alongside these testing strategies to minimize ongoing transmission during the COVID-19 pandemic.^4^

## Data Availability

All data and code publicly available.

## Acknowledgments

Some of the computing for this project was performed on the Nero platform. We would like to thank Stanford University, the Stanford Research Computing Center, and Stanford Medicine for providing computational resources and support that contributed to these research results.

## Authorship contribution

Drs. Nathan C. Lo and Mathew V. Kiang had full access to all of the data in the study and take responsibility for the integrity of the data and the accuracy of the data analysis.

Study design-NCL, MVK

Data analysis-MVK, ETC, BQH, LACC, NCL Data interpretation-All authors

Wrote first draft-NCL

Contributed intellectual material and approved final draft-All authors

## Role of the Funding Organization or Sponsor

The funding organizations had no role in the design and conduct of the study; collection, management, analysis, and interpretation of the data; and preparation, review, or approval of the manuscript; or the decision to submit the manuscript for publication.

## Previous presentations

None.

## Research in context

### Evidence before this study

We searched PubMed for relevant articles published in English before December 7, 2020, using the search terms “travel” or “air*” together with “COVID-19” or “SARS-CoV-2” with “screen*” or “test*” restricted to the title and abstract fields. This search identified 87 articles. We found one article that discussed risk of international importation of infections during the early pandemic.^33^

### Added value of this study

This study provides an analysis of routine testing strategies to reduce risk of COVID-19 associated with airline travel. We find that pre-travel testing strategies, including same-day rapid antigen testing and pre-travel PCR testing (within 3 days prior to departure) reduced the risk of traveling while infectious. Notably, the same-day rapid antigen testing performed slightly better than the pre-travel PCR test when assuming performance characteristics for the rapid antigen test comparable to the Abbott BinaxNow test. We examined the addition of PCR 5 days after arrival and found that this reduced the overall infectious days during the travel period, providing a larger population-level effect through reduced importation of infections. The relative benefit of the post-travel testing was related to the incidence of the destination city, with largest benefit when traveling from a high to low incidence setting.

### Implications of all evidence available

The study findings suggest that test-and-travel strategies for COVID-19 will likely improve the safety of airline travel and can be a public health tool to reduce importation of infections into low incidence settings. Routine testing for COVID-19 prior to travel, including same-day rapid antigen testing, can be an effective strategy to reduce individual risk of COVID-19 infection during travel. However, post-travel testing with abbreviated quarantine is likely needed to reduce population-level transmission from importation of infection, especially for travelers going from a high to low incidence setting. All testing strategies were imperfect, and should be viewed as a tool to be used alongside social distancing, universal masking, and other infection control measures during travel.

## References

1. Fauci AS, Lane HC, Redfield RR. Covid-19 - Navigating the Uncharted. N Engl J Med 2020; 382(13): 1268–9.

2. Zhu N, Zhang D, Wang W, et al. A Novel Coronavirus from Patients with Pneumonia in China, 2019. N Engl J Med 2020; 382(8): 727–33.

3. Chu DK, Akl EA, Duda S, et al. Physical distancing, face masks, and eye protection to prevent person-to-person transmission of SARS-CoV-2 and COVID-19: a systematic review and meta-analysis. Lancet 2020; 395(10242): 1973–87.

4. Lewnard JA, Lo NC. Scientific and ethical basis for social-distancing interventions against COVID-19. Lancet Infect Dis 2020.

5. Hsiang S, Allen D, Annan-Phan S, et al. The effect of large-scale anti-contagion policies on the COVID-19 pandemic. Nature 2020; 584(7820): 262–7.

6. The Lancet Infectious D. Air travel in the time of COVID-19. Lancet Infect Dis 2020; 20(9): 993.

7. Choi EM, Chu DKW, Cheng PKC, et al. In-Flight Transmission of SARS-CoV-2. Emerg Infect Dis 2020; 26(11): 2713–6.

8. TSA. TSA checkpoint travel numbers for 2020 and 2019: https://www.tsa.gov/coronavirus/passenger-throughput?page=1 accessed 12/1/20, 2020.

9. Li R, Pei S, Chen B, et al. Substantial undocumented infection facilitates the rapid dissemination of novel coronavirus (SARS-CoV-2). Science 2020; 368(6490): 489–93.

10. United Airlines Becomes First U.S. Carrier to Make COVID-19 Tests Available to Customers. 2020. https://hub.united.com/2020-09-24-united-airlines-becomes-first-u-s-carrier-to-make-covid-19-tests-available-to-customers-2647787602.html.

11. Hawaii: Coronavirus (COVID-19) transportation related information and resources: https://hidot.hawaii.gov/coronavirus/. 2020 (accessed 11/6/20.

12. CDC. Testing and International Air Travel https://www.cdc.gov/coronavirus/2019-ncov/travelers/testing-air-travel.html accessed 12/1/20, 2020.

13. Chin ET, Huynh BQ, Chapman LAC, Murrill M, Basu B, Lo NC. Frequency of routine testing for COVID-19 in high-risk healthcare environments to reduce outbreaks. Clin Infect Dis 2020.

14. He X, Lau EHY, Wu P, et al. Temporal dynamics in viral shedding and transmissibility of COVID-19. Nat Med 2020; 26(5): 672–5.

15. Khanh NC, Thai PQ, Quach HL, et al. Transmission of SARS-CoV 2 During Long-Haul Flight. Emerg Infect Dis 2020; 26(11): 2617–24.

16. Myers JF, Snyder RE, Porse CC, et al. Identification and Monitoring of International Travelers During the Initial Phase of an Outbreak of COVID-19 - California, February 3-March 17, 2020. MMWR Morb Mortal Wkly Rep 2020; 69(19): 599–602.

17. CDC COVID Data Tracker: https://covid.cdc.gov/covid-data-tracker/#cases_casesper100klast7days 2020 (accessed 11/10/20.

18. Oran DP, Topol EJ. Prevalence of Asymptomatic SARS-CoV-2 Infection: A Narrative Review. Ann Intern Med 2020.

19. Sun K, Wang W, Gao L, et al. Transmission heterogeneities, kinetics, and controllability of SARS-CoV-2. Science 2020.

20. Bajema KL, Wiegand RE, Cuffe K, et al. Estimated SARS-CoV-2 Seroprevalence in the US as of September 2020. JAMA Intern Med 2020.

21. Kiang MV. GiHub Repository: github.com/mkiang/airline_testing_strategies. 2020.

22. CDC. Testing and International Air Travel: https://www.cdc.gov/coronavirus/2019-ncov/travelers/testing-air-travel.html. accessed 11/27/20., 2020.

23. Kucirka LM, Lauer SA, Laeyendecker O, Boon D, Lessler J. Variation in False-Negative Rate of Reverse Transcriptase Polymerase Chain Reaction-Based SARS-CoV-2 Tests by Time Since Exposure. Ann Intern Med 2020.

24. Hay AH, Kennedy-Shaffer L, Kanjilal S, Lipsitch M, Mina M. Estimating epidemiologic dynamics from single cross-sectional viral load distributions. Pre-Print 2020.

25. Gibani MM, Toumazou C, Sohbati M, et al. Assessing a novel, lab-free, point-of-care test for SARS-CoV-2 (CovidNudge): a diagnostic accuracy study. Lancet Microbe 2020; 1(7): e300–e7.

26. Albert E, Torres I, Bueno F, et al. Field evaluation of a rapid antigen test (Panbio™ COVID-19 Ag Rapid Test Device) for the diagnosis of COVID-19 in primary healthcare centers. Pre-Print 2020.

27. Pilarowski G, Lebel P, Sunshine S, et al. Performance characteristics of a rapid SARS-CoV-2 antigen detection assay at a public plaza testing site in San Francisco. Pre-Print 2020.

28. Schwob JM, Miauton A, Petrovic D, et al. Antigen rapid tests, nasopharyngeal PCR and saliva PCR to detect SARS-CoV-2: a prospective comparative clinical trial: https://www.medrxiv.org/content/10.1101/2020.11.23.20237057v1.full.pdf. 2020.

29. Grassly NC, Pons-Salort M, Parker EPK, White PJ, Ferguson NM, Imperial College C-RT. Comparison of molecular testing strategies for COVID-19 control: a mathematical modelling study. Lancet Infect Dis 2020.

30. Paltiel AD, Zheng A, Walensky RP. Assessment of SARS-CoV-2 Screening Strategies to Permit the Safe Reopening of College Campuses in the United States. JAMA Netw Open 2020; 3(7): e2016818.

31. Larremore DB, Wilder B, Lester E, et al. Test sensitivity is secondary to frequency and turnaround time for COVID-19 screening. Sci Adv 2020.

32. Lauer SA, Grantz KH, Bi Q, et al. The Incubation Period of Coronavirus Disease 2019 (COVID-19) From Publicly Reported Confirmed Cases: Estimation and Application. Ann Intern Med 2020; 172(9): 577–82.

33. Dickens BL, Koo JR, Lim JT, et al. Strategies at points of entry to reduce importation risk of COVID-19 cases and re-open travel. J Travel Med 2020.

